# Descriptive and Narrative Study of Long Covid Cases in General Practice and Diagnostic Value of Single Photon Emission Computed Tomography (SPECT scan)

**DOI:** 10.1101/2022.03.01.22270897

**Authors:** Marc Jamoulle, Gisèle Kazeneza-Mugisha, Ayoub Zayane

## Abstract

Primary care is under great pressure from patients with Covid -19 and those affected by Long Covid. The issue of Long Covid, its diagnosis and therapeutic approach are discussed here in detail. The Long Covid is described on the basis of a review of the literature and also on the basis of clinical experience in general practice. The main characteristics of thirty four cases (twenty five women) of Long Covid encountered in 2021 and early 2022 are outlined. The experience of six of them is reported on the basis of notes from their medical records. These six patients were interviewed and each was asked to reread and correct the texts concerning them. This is therefore a descriptive study based on clinical and narrative experience, verified by the patients.

Long Covid, the first disease in the history of medicine to be described first by patients themselves on social networks, is not yet precisely defined and the multi -systemic symptoms may be non-specific or vary according to the organs affected.

Diagnosis is based on careful listening to the patient’s history. Previously unknown irrepressible fatigue, brain fog, working memory disorders with possible anomia, anosmia, dysgeusia or other muli-systemic symptoms occurring after an acute Covid are varying characteristics of Long Covid. Biological evidence of Covid is missing in fourteen patients as PCRs may have been not done or came back negative in the acute phase of the disease. Anti-SARS-CoV-2 antibodies are not always present or are indistinguishable from post-vaccine antibodies. In fourteen severe cases presented, Single Photon Emission Computed Tomography (SPECT scan) after intravenous administration of Technetium-99m (Tc-99m HMPAO) were able to demonstrate a disorder of cerebral perfusion. Two follow -up brain SPECT at three months showed significant improvement. Further genetic and immunologic study is ongoing for all patient with the help of the international consortium COVID Human Genetic Effort. A patient who presents after a Covid with medically unexplained symptoms may well be a Long Covid. Despite some interesting hypothesis, there is no known specific treatment. Neurocognitive revalidation and physiotherapy may help those patients who need long -term empathic support to cope with their condition.

**Key messages:**

□ Long Covid is a recent onset, multi-systemic, long-term condition that can be very debilitating.
□ The main symptoms are severe fatigue, exertional exhaustion, and cognitive and memory problems, among others.
□ Patients who suffer from it may not realize it, may not talk about it, or may attribute their problem to other causes.
□ Single Photon Emission Computed Tomography (SPECT CT) contributes to the hypothesis of a vascular perfusion disorder induced by SARS -coV-2 and should be validated as a diagnostic tool in neurological Long Covid.
□ Tissue immunity should be available to prove Long Covid in case of humoral seronegativity
□ There is no identified treatment that can be recommended yet. Careful listening, empathic support and cognitive and physical rehabilitation are suggested and should be organised or supported by the Belgian state.

## 1 Introduction; Long Covid in family medicine

Covid-19 is no longer just an acute disease. In a large number of previously infected patients, which, depending on the authors, can vary from 10% to more than 50%, the disease is no longer just an acute disease.^1, 2^ It can develop into a disabling disease, sometimes lasting several months, called Long Covid, which only appeared in the literature in the summer of 2020.^3, 4^ Diagnosis is still difficult, as the signs and symptoms have untested diagnostic properties. The Long Covid is also very difficult to define^5^ and a Delphi consensus, led by the WHO, was needed to reach a definition.^6^ The vision of Long Covid in the international literature still has a strong selection bias. The articles that deal with it are very hospital-centric and talk more about the doctor’s view on Long Covid than the patient’s.^7^

It is a multi-system disease, most often occurring after a relatively mild acute illness^8^ or an influenza-like illness, and encompasses distinct groups of heterogeneous symptoms that may overlap, evolve over time and are sometimes difficult to link to Covid-19. It is also the first disease to be named by patients themselves through exchanges on social networks. Patient advocacy groups, many of whose members identify themselves as Covid Long haulers, are also mobilizing via the Internet and social networks, for example in France : <u>(apresj20.fr),</u> in the United Kingdom : (longcovidsos.org), in the Netherlands : <u>(coronaplein.nu),</u> in Germany : (longCoviddeutschland.org), Canada :(covid-longhaulcanada.com) or United States of America : (survivorcorps.com). In the Netherlands, a site launched for the RECOVER study^9^ allows patients to do a self-test <u>(Moe na corona?</u>) Numerous support groups for Covid long have appeared on Facebook, with tens of thousands of members. All these patients willingly participate in online surveys.

Long Covid syndrome occurs in patients who often already have a high level of comorbidity.^10^ They also sometimes find it difficult to be heard and may be labelled as *heartsink patients* (unbearable patients).^11^ The disease can be very debilitating and is becoming a serious public health problem, with almost one million people affected in the UK alone.^12^ The term *Long Covid* refers to symptoms persisting for more than four weeks after the diagnostic, usually managed in general practice. Unbearable fatigue, brain fog^13^ and myalgia are the most common symptoms. Symptoms can be multiple and of varying intensity depending on the areas of the body affected. Cognitive disorders, memory and attention deficits, anomia, dysarthria, frontal behavioral disorders, autonomic dysregulation, headaches, dyspnea, anosmia, dysgeusia, skin or digestive disorders, psychosocial distress, loneliness, anxiety, depression and sleep disorders have been associated with Long Covid.^14, 15, 16,^ 17

A distinction must be made between (1) post-Covid patients whose general condition has been severely impaired after cardiac, pulmonary or other damage as a result of a severe Covid, and (2) those who a few months after a Covid that may have seemed trivial still present with an unusual syndrome of exhaustion, neurocognitive or mental disorders with memory impairment and in particular word loss. A meta-analysis shows that after an acute episode with hospitalization, the most frequent symptoms are chest pain, fatigue, dyspnea and cough.^18^ In Long Covid, exhaustion, memory and concentration problems are at the forefront and the quality of life is very impaired.^19^

The absence of specific markers means that the diagnosis is based on the patient’s word, which is not without medico-legal consequences.^20^ Positron-Emission Tomography (PET scan) could provide precise information such as the hypo-metabolism that affects certain brain areas in certain Long Covid with a strong neurological component.^21, 15, 22, 23^ Reversibility of the decrease in neocortical 21 glucose metabolism assessed by 2-[18]-Fluoro-2-deoxy-D-glucose positron emission tomography, accompanied by an improvement in cognitive function, may occur after 6 months in some patients.^24^ However, this procedure is not easily accessible and is also expensive. Single photon emission computed tomography (SPECT scan), on the other hand, are more accessible and may reveal a metabolic brain disorder similar to that found in Alzheimer’s disease or stroke.25

To date, we haven’t reached a full understanding of what Long Covid really is, nor its natural history.^26^ There is no evidence of its prevalence or pathology and there is no specific treatment.^27^ Moreover, although some research seems encouraging, it is not clear that vaccination protects against Long Covid.^28^ Family doctors can be valuable sources of explanation. The doctor can certainly provide information and support to these distressed and sometimes stigmatized patients.^29^ Health inequalities are the norm in many countries and as Berger et al state, “*primary care providers are in a unique position to provide and coordinate care for vulnerable patients with post-Covid syndrome*”.^30^ Psychotherapeutic care, neurocognitive revalidation and physiotherapy seem to provide some comfort^31, 32^ although treatments are still controversial. Parkin et al. propose “*an integrated, multidisciplinary model of care to deal with the growing number of cases*”.^33^ This formulation makes an interesting reference to the descriptions of the family doctor’s work^34^ which could very well, as Ward et al. point out, become *the manager of such an approach*.^35^ There are as yet no definitive, evidence-based recommendations for the management of the Long Covid. Patients should therefore be managed pragmatically and symptomatically.^36^ A study by Catalan et al. suggests that patients who receive corticosteroids on admission for acute hypoxemic Covid-19 are less likely to have persistent symptoms and more likely to have a better quality of life at one year after admission.^37^

When the usual biological examinations do not explain the fatigue and do not lead to a diagnosis, the patient’s word remains. Narrative medicine,^38^ is based on trust, expressed in a consultation between two people who have often known each other for a long time. Relational continuity^39^ is one of the characteristics of general practice, and this continuity enables the family doctor to cope with polymorbidity and its complexity.^40^ One third of patients with post-Covid syndrome have pre-existing comorbidities.^10^

The family doctor^a^ is a natural interlocutor and will therefore be well positioned to exercise his ability to find a problem^41^ which is then turned into a research question. It is about understanding how Covid-19 is responsible for unexpected symptoms which overwhelm the patients, their families and their employers, who may not understand the situation. This empathetic relationship also allows the patients to state complaints that are not acceptable to themselves, such as exhausting fatigue or a previously unknown feeling of depression, which they will not dare to express to their family and makes them fear the future.^42^ Studies on workers showed that many do not consult their doctor about the syndrome and face social stigma when returning to work.^43, 44^

In his/her practice, it is paramount for the family doctors to take a careful history of the patient, to ask about previous infection with Covid-19 and to detail and record all symptoms and their timing, ranging from mild to severe symptoms that may affect all organs. It is also essential to assess cognitive impairment and psychological impact.^45^ A GP should not be surprised to hear a patient with Long Covid saying: “*My body and I go our separate ways*”.

The certainty of the diagnosis itself is low and is primarily clinical. Therefore, a careful history will be essential to support the patient, to identify and “*name*”^46^ this tiring disease which seems endless and without remedy. Moreover, the patient may on the one hand feel misunderstood and neglected, and on the other hand not get the appropriate health care that she/he needs. In turn, a doctor may feel that he or she has failed by being reduced to listening and comforting when that is precisely its role. If the doctor does not identify the problem and diagnoses a case of medically unexplained symptoms, depression or an anxiety attack, the patient will feel unrecognized in his suffering and will lose confidence in the doctor. The relationship therefore cannot be established. This is a typical case of the application of quaternary prevention whose aim is to strengthen the doctor-patient relationship and thus avoid harm.^47^

In order to contribute to the knowledge of Long Covid, we believe that the voices of patients are essential. The aim of this publication is to give patients a voice, to identify their problems and to understand how we can best support them. Clinical medicine tries to integrate the experience of the clinician, the values of the patient and the best scientific information available. These three pillars are also the pillars of Evidence Based Medicine.^48^ But in this case, there is no evidence yet, it is under construction as patients and doctors work together to develop knowledge. It is therefore a type of action research^49^ by which, as J. Pols points out, *it is up to us to articulate the knowledge that patients develop*.^50^

## 2 Method; long-term follow-up and patients involvement

### 2.1 The consultation as a place for research

The setting is that of a general practice in Belgium, in a group practice of family doctors working on a fee-for-service basis in a long-term relationship of trust with their patients, based on interpersonal continuity. The general practitioner, for whom the Covid load has changed the professional practice by making it more accessible, in particular through remote consultation,^51^ is the patient information manager. As such, he ensures informational continuity^52^ by keeping a diagnostic index (list of problems) and a therapeutic index (list of medicines) as well as a daily journal. The GP also has a research and teaching function and receives students on placements. It should be taken into account that each consultation can initiate a research process.

### 2.2 A summary table of cases available online

Patients seen since the beginning of the epidemic and who have been considered for a Long Covid are listed since early 2021 anonymously in a summary table available online with some clinical and biological characteristics. This online Google Sheets file, available in the “Results “section, includes the following elements if available:

– General : ID,
– Dates : acute Covid, Long Covid diagnosis, positive PCR if any, Covid humoral serology if any, 2nd vaccination, scintigraphy, CT-Scan or NMR if any, DUSOI, COOP charts
– Symptoms: acute Covid, Long Covid.
– Certainty (yes/no) of clinical laboratory.
– Assessment scales : overall severity by Dusoi/WONCA, functional status by COOP/WONCA charts
– Protocols : scintigraphy, CT-Scan or NMR

The table is available online: https://tinyurl.com/tablelongcovid). It is also intended to be used for further research into the genetic and immunological profiles of the patients as part of an international research effort by the European Genetic Consortium.53 The online file has 2 sheets. The first one lists the twenty one cases considered as relevant with their characteristics. The second explains the name of the fields. Gender, age, co-morbidity, and other more personal details have been removed from this table in the interests of confidentiality.

### 2.3 Clinical research tools

#### 2.3.1 Description of co-morbidity

Clinically important health problems experienced by patients are indicated by means of the alphanumeric codes of the International Classification of Primary Care 2nd edition (ICPC-2)^54^ already used in the Electronic Medical Records (EMR) in Belgium. These codes has been removed from the online sheet in the interests of confidentiality but are available for specific studies. ICPC-Codes are available on https://www.hetop.eu/hetop/3CGP where the reader can find their detailed description in many languages.^55^ A double A4 page containing all the headings of ICPC-2 is available online in several translations.^56^

#### 2.3.2 Certainty

Clinical certainty is considered here, i.e. the conviction by the practitioner that the anamnestic and paraclinical elements at his/her disposal allow him/ her to reject with a high probability that the case presented is due to another condition.^57^ In clinical practice, the decision must be made, even if the practitioner knows that the statement still contains a possibility of uncertainty. Only cases deemed clinically certain are displayed in the first sheet of the table. Biological certainty, also noted as “yes” or “no”, refers here to the availability of a positive PCR test and/or positive Covid serology before vaccination. At present, only humoral immunity is available.

#### 2.3.3 Severity

The degree of severity is assessed by the doctor using the Duke Severity of Illness Checklist (DUSOI) adapted to family medicine under the name DUSOI/WONCA.^58, 59^ The index is based on four parameters: symptom status, complications, prognosis over the next six months without treatment, and treatability, i.e. the expected response to treatment. The score ranges from 0 (no severity) to 5 (extremely severe). The DUSOI/WONCA form with instructions for use is available online.^60^

#### 2.3.4 Functional status

The achievement of functional status is assessed by the patients themselves using the six COOP/WONCA charts.^61^ General condition, ability to perform daily activities, physical condition, emotional management, ability to have a social life and changes in health status are indicated on the 6 corresponding COOP/WONCA charts by patients rating themselves from 1 (good performance) to 5 (cannot do it). The overall index will have a value from 6 (excellent) to 30 (totally impaired functional status). A manual with the 6 COOP/WONCA cards in several languages and instructions for use is available online.61

### 2.4 Narrative medicine and qualitative approach

A detailed clinical medical case report was written by the doctor for all patients. Six patients seen who showed highly significant signs of Long Covid were visited by GK, a medical student who was not involved in the care. According to the principles of narrative medicine,38 GK conducted the interviews on the basis of standardized semi-open questions. These recorded interviews were transcribed in full and will be used for further qualitative research. GK then reviewed and corrected if necessary the clinical case reports with each six patients.

### 2.5 Laboratory and imaging

The results of PCR tests and Covid humoral serology are indicated with the dates of their performance when available. Sars -Cov 2 serology is not routinely requested in Belgium, as there is a charge for it. None of the other usual laboratory tests requested during the development process proved to be contributory. They are therefore not mentioned in the table.

Single Photon Emission Computed Tomography (SPECT CT)(ECD Tc-99m), an old technique,62 allows the assessment of brain metabolism using complex technetium -99m (Tc-99m) labeled molecules that are able to cross the blood - brain barrier. The binding of these tracers is dependent on cerebral blood flow. Tc-99m is used for brain perfusion studies because of its high first-pass extraction fraction and high affinity for the brain.63

When the are aappears in red, it means that it is working properly. This examination was requested in patients with a severely impaired functional status. The available protocols are reproduced in the table and figures below. Brain CT scans and Nuclear Magnetic Resonance scans were also collected

### 2.6 Confidentiality and ethics

The patients have often been known to the providers for a long time and have formally accepted the management of their clinical information in the framework of the Belgian computerized medical file called *‘Dossier Médical Global*’(Comprehensive Medical Record). Each patient interviewed gave formal written consent to use their data and to publish their story under a pseudonym. The ethics committee of the University Hospital of Liège, Belgium, gave its full approval to this study under the number 2022/23.

### 2.7 Bibliographic follow-up

In parallel with the clinical follow-up, a bibliographic watch on PubMed and Google Scholar on the subject of Long Covid allowed us to address the complexity of this new situation. The following MeSH descriptors “post-acute Covid-19 syndrome” [Supplementary Concept] OR “Long Covid” [TW] were incorporated into the PubMed alert system.

## 3 Results

### 3.1 Overall description of thirty-four cases monitored in 2021 and early 2022

All cases of Long Covid encountered and followed up at the Janson Medical Centre during the year 2021 and early 2022 have been analyzed and their main characteristics are presented in a summary table accessible online: https://tinyurl.com/tablelongcovid

#### Data from thirty-four patients

Between May 2021 and February 2022, thirty -four patients, from age range 15-19 to 60-64 years, of whom sixteen women, were clinically identified as suffering from Long Covid (Table 1. The average age of these patients is 40 years. PCRs were positive in only ten of the 21 patients. The other PCRS have been either not done or negative. Ten patients have a professional occupation.

**Table 1.** Patients with clinical Long Covid met in general practice. Data collected from May 2021 to February 2022. See also: https://tinyurl.com/tablelongcovid

|  |  |
| --- | --- |
| Number of patients with clinical Long Covid | 34 patients |
| Number of women | 25 (73%) |
| Mean age | 40 |
| Number of patients with no laboratory proof | 14 patients |
| Number of positive PCRs | 20 patients |
| Number of cerebral Tc-99m HM-PAO with altered perfusion | 14/14 ordered |
| Number of patients severe to very severe (DUSOI 3 or 4) | 31/34 patients |
| Number of patients with altered to very altered functional status (score more than 22 on COOP /WONCA Charts if available | 20/23 patients |
| Mean number of months before diagnosis | 12 (min 1 max 20) |
| Mean duration of the Long Covid (months) | 10 (min 2 max 20) |
| Number of days off work for 9 patients | ~1275 (30 to 350) |

At the end of 2021, two patients were still fully off work, six were working part-time. There were approximately 1275 days off work since the onset of the Covid disease for these eight people. For two patients, the illness was contracted at work and an accident at work report was submitted.

The comorbidity, indicated by ICPC-2 codes, is for some, very heavy (polyarthritis (L88), diabetes (T90), asthma (R96), common migraine (N89), major obesity followed by partial gastrectomy (T83), Meniere syndrome (H82), a miscarriage during acute Covid (W82), a dissecting carotid aneurysm during Covid (K99), a transient ischemic attack (K92), both due to or during acute Covid.

Five patients contracted Covid at the very beginning of the epidemic in early 2020. Six patient had acute Covid twice. Two patients improved following the vaccine while one worsened. Two patients experienced serious side effects following vaccination. One patient was hospitalized for acute Covid. Two patients had home oxygen. All but one patient were vaccinated after having been ill with Covid.

The symptoms of Covid are typical. The symptoms experienced during Long Covid are numerous, as are the variations between patients. The previously absent disabling fatigue, inability to exert oneself and brain fog with memory impairment and loss of speech form a recurrent picture. Covid serology, when available, were most often performed after vaccination.

The identification of Long Covid disease took between 1 and 19 months. The patients who consulted were given a wide range of diagnoses. A school-girl was called a lazy teenager by her teacher. Angor, pulmonary embolism, multiple sclerosis, depression, fibromyalgia, burnout, generalized anxiety disorder or post-traumatic shock were reported in the emergency or specialist reports.

#### Single Photon Emission Computed Tomography contribution

Single Photon Emission Computed Tomography (SPECT CT) were requested in fourteen patients with worrying indicators and neurological status. In one reported case the SPECT CT was not requested due to a desire to become pregnant. The fourteen SPECT CT requested, five of which are documented below (see Figure 2 to Figure 6), all showed altered cerebral perfusion, sometimes extensive. In a case of Long Covid without memory impairment, there is no impairment of thalamic or sub thalamic perfusion. In two cases, a control SPECT CT, requested after 3 months, showed an improvement in cerebral perfusion, well in parallel with the clinical improvement of these patients. Nuclear magnetic resonance (NMR) are generally non-contributory except for mere minimal lesions. A follow -up NMR showed normalization of an aneurysmal lesion present in the patient during Covid.

**Figure 1.**
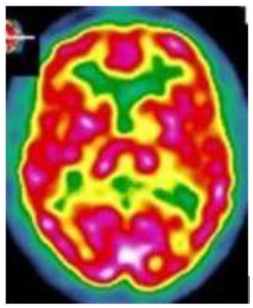
Normal Single Photon Emission Computer Tomography. Courtesy; A. Kas, APHP, Paris

**Figure 2.**
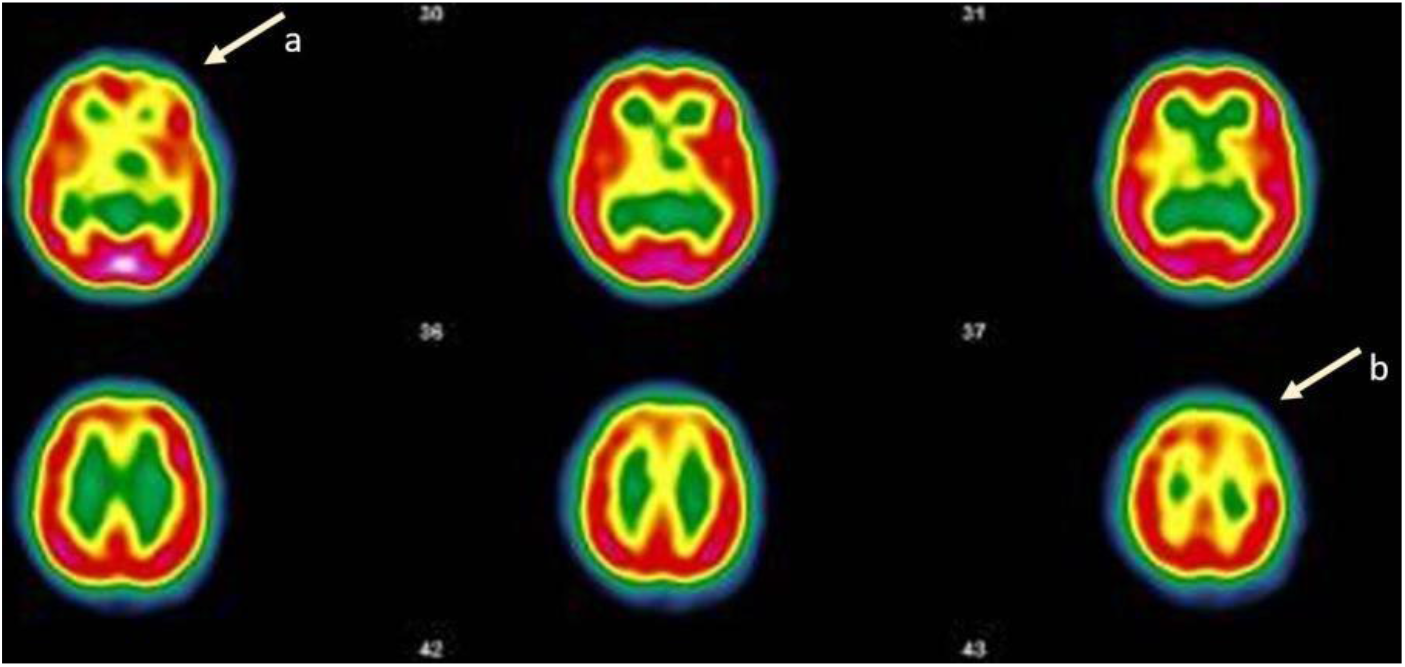
Case 1; 2021 On the images taken, left frontal (a), left fronto-parietal (b) and left thalamic hypofixation is seen. The fixation in front of the cerebellum is correct. Conclusion : Scintigraphic examination (SPECT CT) (ECD Tc-99m) compatible with a cerebral pathology of the vascular type with clearer left fronto -parietal, left frontal and left thalamicvascular disorders (Images and protocol : Drs Bouazza and Mahy, Vesalius Hospital, ISPPC, Belgium)

#### Patients with severely impaired health

The severity of the cases, as estimated by the doctors using the DUSOI, shows very severe impairment (rated 4) in 15 patients and severe impairment (rated 3) in 12 patients. Functional impairment is very high (scored more than 20 on the six WONCA /COOP cards) in twenty of the twenty three patients tested.

### 3.2 Narration of six exemplary cases

The cases reported below have been reviewed and approved by the patients.

#### Case n° 1

A single mother of two children, age range 40 -49, without many resources and dependent on social support, has been a patient of our practice for several years. In late 2020, she had a severe Covid for 3 weeks at home with dyspnea, sore throat, no fever, dysgeusia and a positive PCR test for SARS-CoV-2. Nine months later, she had still not recovered.

Although the patient was already known to have fatigue due to thalassemia minor and occasional depressive feelings, the history of an acute Covid, with in addition, severe and very disabling fatigue, sleep disturbance, headaches, dysgeusia, unusual and more frequent depressive feelings, significant cognitive problems, including omissions and partial anomia (loss of word) placed her firmly in the Long Covid category. The last consultation, a fortnight earlier, had been difficult. She had explained that she was crying a lot, that her nights were haunted by strange hallucinatory dreams and that she could no longer cope.

Completely unexpectedly, this patient was much improved by the Comirnaty vaccine. The vaccine injections made her extremely ill to the point of bed rest but she then felt almost cured. Most of the long lasting symptoms have disappeared, the only ones still present are headaches and some memory problems.

There is a minimal abnormality showing on the NMR. The usual laboratory tests showed no alterations. The anti-SARS-CoV-2 antibody level was very high (800.0 AU /mL). Three weeks later, the SPECT (Tc -99 m DCE) was consistent with a vascular type of brain pathology (see figure 2).

Unfortunately, after 6 months, in early 2022, this patient suffered a severe relapse with very migraine, hallucinatory dreams and recurrence of fatigue. The effect of the vaccination was therefore transient. A case report had already been proposed for publication but will be revised due to the evolution of the disease.64

#### Cas n° 2

A women, age range 35-39, had Covid in early 2020. Headache, persistent dry cough (does not smoke), joint and muscle pain (like flu), intense fatigue and bed rest; no fever, no anxiety, no depressive element. Although she was concerned about many her symptoms, she expressed real concerns regarding (1) chest pains that were more intense than the intercostal pain (that did not vary with breathing) she had experienced before, and (2) her additional breathing difficulties. After about ten days, she then recovered from these additional breathing difficulties.

Three months later, she had a choking sensation at night; she had to control her breathing with yoga and visualization techniques (meditation techniques) and eventually fell asleep. The suffocating sensation remained, gradually manifested itself even during the day, but it did not prevent physical effort. This was followed by symptoms such as neck pain, arm pain and paresthesia in the fingers, which also disappeared within a few days.

In August-September 2020: ten days of intense headaches, similar to those she was already prone to, but persisting longer than usual and with permanent jaw pain. Her GP could not explain the headaches, and feared a neurological problem; unremarkable bio. The headaches went away as they came. In the meantime, the breathing problems have not disappeared.

In early 2021, she had another asymptomatic Covid, with positive PCR. During this period, she suffered a miscarriage and a blood test carried out in this context showed that the patient did not have any anti -SARS -CoV -2 antibodies.

In the spring of 2021, supposedly the beginning of her Long Covid, she is drained, tired, with renewed chest pain, respiratory stress, recurrence of ulnar pain, sciatica -like pain in the left leg, left hypochondrium with pain on breathing, accompanied by paresthesia in the digital area and pain in the left arm, intense fatigue, disturbed sleep, headaches, profuse sweating; and feels unable to work due to her symptoms, exacerbated by anxiety. Two months later is the beginning of a dark period with intense panic and irrational behavior (fear of fainting in the bath). She uses breathing control techniques; she cannot stay alone (for the first time in her life), manages to get help and activates her network of friends. She no longer recognizes herself. She still has a lot of emotional moments and breathing mechanics problems.

In summer 2021: three weeks during which the patient feels a considerable improvement. During this month, she had a vaccination against Covid-19, and felt as if she had been cured afterwards. She regained physical shape with a sports coach : no more anxiety attacks, the brain fog and dizziness improved and almost disappeared, but she still experienced unusually excessive tiredness.

The chest pain returned in autumn as well as the fatigue. However, her fatigue can also be attributed to the fact that the patient has been pregnant since mid-August. Since then, the patient says that she is worn out by her pain and feels that it will not go away. At present, the chest pain seems to have subsided, but the headaches have returned. The desire to become pregnant has led to her not requesting a SPECT.

#### Case n° 3

A age range 45 -49 old asthmatic patient suffering from Meniere ‘s syndrome with severe vertigo and from common disabling migraines. In late 2020, Covid treated at home with pneumonia (ground glass areas on the X-ray): fatigue, dyspnea at the slightest effort, chest pain and anosmia. After one month, the patient writes this text message: “*For the past few weeks, I have been having obsessive and fixed ideas, I have experienced paranoia about my colleagues, violent anxiety and sometimes morbid ideas with a consequent state of despair, crying spells that cause chest pain, recurrent nightmares, and I sometimes have the feeling of being in a waking sleep where I am aware of having very strong chest pains, bones and joints* … *In the morning, I have no strength, no motivation, I feel recluse and even persecuted*… *There is something wrong*… “.

Subsequently, the chest pain persists and her dizziness worsens, to the point that she has had twelve dizzy spells in just forty days. She finds it difficult to concentrate, she can’t stand noise, and she loses her words and her memory. She is exhausted and develops sleep disorders. It is the SPECT CT the key. There are severe cerebral vascular lesions (see figure 2). The same patient, whose condition gradually improved, had a second SPECT CT three months later. The protocol of this second SPEC<u>T</u> is very reassuring: a discreetly heterogeneous tracer fixation is observed, with clearer left frontal, left parietal and right parietal hypofixations, and the presence of periventricular hypocaptation. Compared to the previous workup, there is an improvement in cerebral fixation with a decrease in fixation heterogeneity and periventricular hypocaptation.

#### Case n° 4

A mother, age range 55-59, and surface technician, currently on disability for osteoarthritis, is not sure of what happened to her. She had had Covid with headaches, asthenia, muscle and joint pain as well as anosmia and agueusia, the latter two symptoms having persisted for 3 months. Now she sometimes has difficulty pronouncing certain words, so much so that she’s stopped speaking. There are also problems with her writing abilities.

She has always been a bit dysgraphic, but now she has lost confidence in herself and her knowledge, she is afraid to make mistakes. She has problems with balance and concentration, occasional coughing fits, sometimes tightness of breathing and dyspnea on exertion. Her fatigue persists and prevents her from doing her work, she feels weak and anxious. She also has memory loss : she forgets where she puts her keys and sometimes loses her words. The SPECT CT shows a small but significant brain damage (see figure 4)

**Figure 3.**
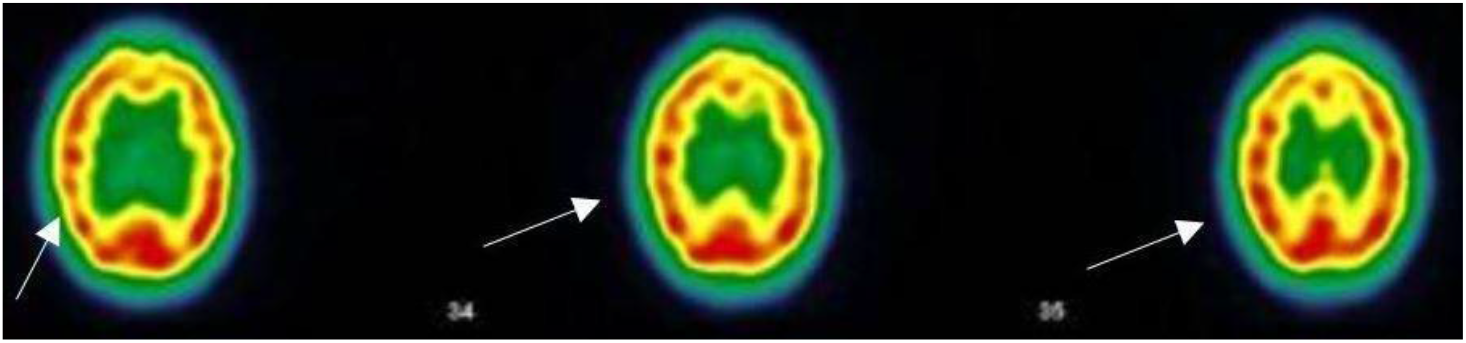
Case n ° 3; SPECT (ECD Tc-99m) : heterogeneous tracer fixation with clearer left frontal, left parietal and right parietal hypofixations. No preservation of the sensory -motor cortices. The fixation in front of the grey nuclei and the cerebellum is correct. Presence of periventricular hypocaptation. Conclusion : Evidence of heterogeneous tracer fixation and periventricular hypocaptation compatible with vascular -type cerebral damage. Three months later, a control examination (not shown here): improvement of the cerebral fixation with decrease of the heterogeneity of fixation and of the periventricular hypocaptation (Images and protocol : Drs Bouazza and Mahy, Vesalius Hospital, ISPPC, Belgium)

**Figure 4.**
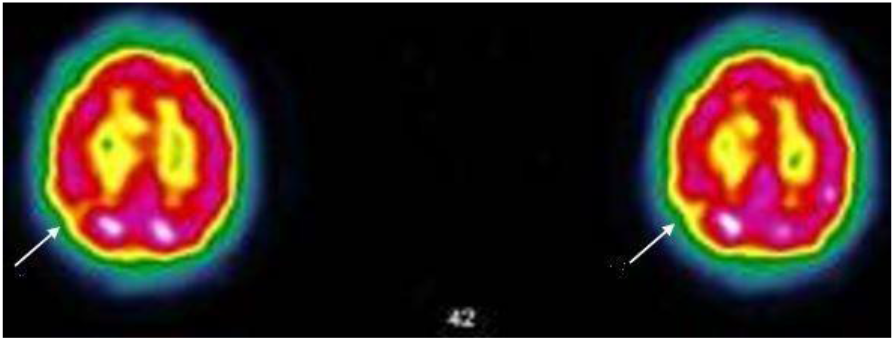
Case n°4; SPECT CT (ECD Tc-99m)) ; heterogeneous tracer fixation with clearer hypofixations left frontal, left parietal, right parietal. No preservation of the sensory-motor cortices. The fixation in front of the grey nuclei and the cerebellum is correct. Presence of periventricular hypocaptation. Conclusion: Evidence of heterogeneous tracer fixation and periventricular hypocaptation compatible with vascular-type cerebral damage. (Images and protocol; Drs Bouazza & Mahy, Vésale Hospital, ISPPC, Belgium

**Figure 5.**
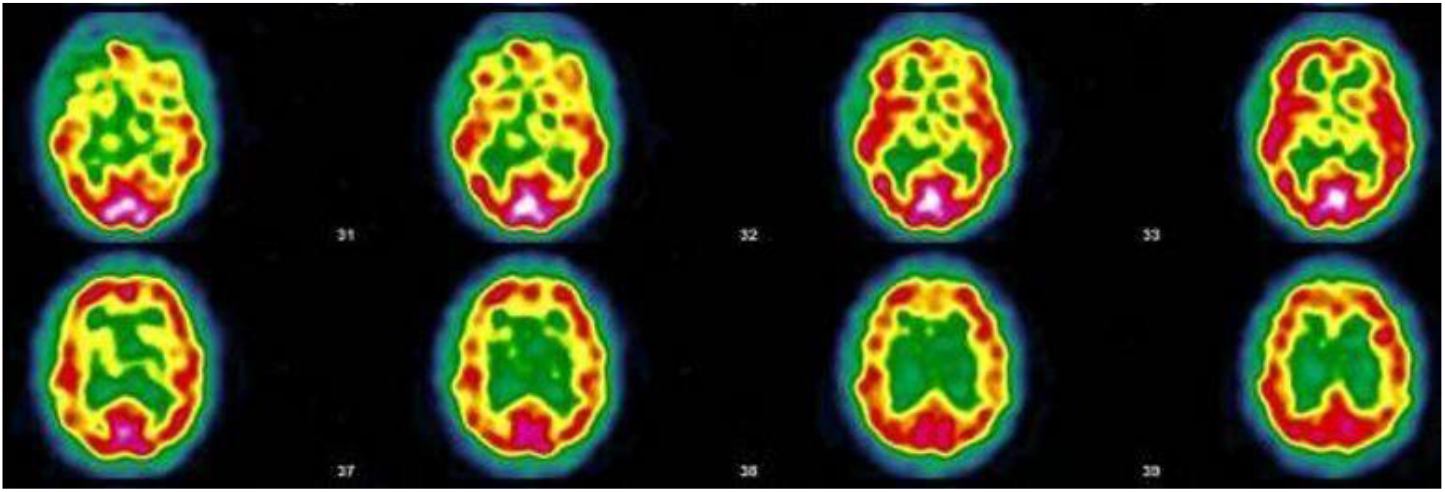
Case n°5; SPECT CT (ECD Tc-99m) ; heterogeneous tracer fixation with bilateral temporal, bilateral frontal, left posterior parietal, right parieto-occipito-temporal hypofixation. Discrete preservation of the sensory-motor cortices. The fixation in front of the grey nuclei is correct. Right cerebellar hypofixation. Cortico-subcortical atrophy with periventricular hypocaptation as an indirect sign. Conclusion: Scintigraphic examination compatible with a cerebral pathology of vascular type. Moderate cortico-subcortical atrophy. (Images and protocol; Drs Bouazza & Mahy, Vesale hospital, ISPPC, Belgium)

#### Case n° 5

A clerk, age range 50-54, in a large hospital. He is being treated for rheumatoid arthritis and brachialgia on foraminal stenosis.

In early 2020 he fell ill with a pseudo flu, which turned out to be a characteristic acute Covid. But at that time, PCR was not yet performed. Several months later, he presented with a characteristic Long Covid with anxious depression, disabling headaches, exertional exhaustion, chest and muscle pain, paresthesia, visual disturbances, nervousness, eye burning, gastrointestinal disturbances, malaise, and above all, a major worry regarding his future.

The patient no longer feels like himself. The SARS -CoV -2 serology is negative. There is therefore no evidence of an occupational disease. The brain scintigraphy shows the characteristics of encephalopathy. At the end of 2021, he was still off work and his functional state was very impaired.

A claim for recognition of an accident at work was made impossible by the absence of a positive PCR test (see figure 5).

#### Case n° 6

A self -employed man, age range 35 -39, in the process of setting up a business, very active, athletic, a jogger and with no previous medical history. In late 2020, on his way home from work, he suddenly feels unwell with a loss of strength. In the weeks that follow, he feels like a stranger to himself and to others.

For several months, he is prostrate on his chair, with some breathing difficulties. The PCR is negative and the blood test, which is ordinary, does not show any SARS-CoV-2 antibodies. He is increasingly depressed, anxious, distressed, atonic, and totally dependent on his family. He is suddenly impaired to the point of not being able to concentrate, feeling totally exhausted, unable jog every day, and is subject to joint and muscle pain. His memory seems impaired and he suffers from hypersialorhoea and hyperhidrosis.

Suddenly, he does not tolerate tobacco or alcohol anymore and stops smoking and drinking. His reactivity to cannabis has also changed. He has become very sensitive to his own cannabis, which makes his heart race even with plants that have only 5%of THC; his fatigue is increasing, he can’t even ride his bike for 10 minutes, has trouble holding the shower over him and catching his breath. He thinks, and so does his doctor, that he’s suffering from a burnout, but neither antidepressants nor sessions with a psychologist seem to help. After a period of improvement, his condition seems to be stationary since early 2021. He experiences dizziness, has to stop in the car, his heart beats too fast, perception of unusual extrasystoles, impression of having drunk, blurred vision, brain fog, headaches, paresthesia in the left arm and jaw. His left thumb has involuntary movements. SARS-CoV-2 serology is one more time negative. In autumn 2021, the brain scan revealed severe vascular damage (see figure 6)

**Figure 6.**
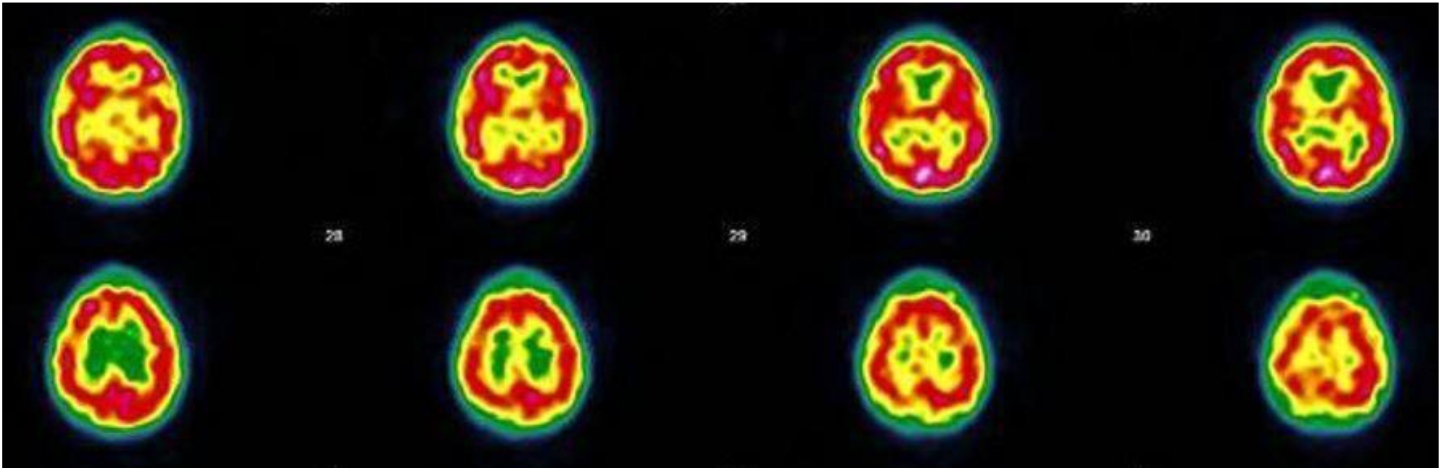
Case n°6; SPECT (ECD Tc-99m)) of ; heterogeneous tracer fixation with more marked hypofixation in the right fronto -parietal, right parieto -occipital, left posterior parietal. No preservation of the sensory motor cortices. Fixation opposite the grey nuclei and cerebellum is correct. Periventricular hypocaptation hypocaptation. Conclusion : Scintigraphic examination compatible with a cerebral pathology of vascular type with more marked right fronto -parietal, right parieto -occipital, left posterior parietal posterior left parietal. (Images and protocol; Drs Bouazza & Mahy, Vesale hospital, ISPPC, Belgium)

## 4 Discussion; an unknown territory

The present study reports thirty-four cases of patients experiencing Long Covid. The general context is that of the Covid-19 pandemic. These cases were progressively highlighted in general practice consultations during the year 2021 and the beginning of 2022. Little by little, the notion of Long Covid imposed itself as a coherent explanation in front of severely altered functional states in known patients. The initial questioning was made during contacts with an abnormally tired patient with impaired memory who suddenly improved after two Comirnaty vaccines. This was the first case (see figure 2) presented above. The remaining patients were then identified one after the other during general practice consultations at our primary health care center.

The diagnosis is therefore essentially made with a clinical narrative approach,^38^ which is based on carefully listening to the patients on multiple occasions. The relationship with most patients had been established for many years, and it was evident that the patients were undergoing a profound change in their condition. The decision to do this observation came directly from clinical experience and the need to understand. This study has therefore a mixed-method approach, with both a quantitative component, the data of which are available in a summary table and a qualitative component.

Regarding the qualitative approach, the first six patients encountered in practice, whose condition was highly challenging, were interviewed by one of the authors (GK). The patients gave their opinions on their own clinical case report written by their doctor and their full interviews will be the subject of a future detailed qualitative analysis.

### Results of the literature review

This study would not have been possible without the availability of biblio-graphic alert systems provided by the National Library of Medicine or Google Scholar. The evolution of the number of publications available on Long Covid has been exponential. Each week brings a new batch of publications. It is the availability of these scientific observations that has allowed us to identify problems, explain them to patients and, sometimes, reassure them. The PubMed interface offers 59 citations in 2020, and 758 in 2021 as of 10 December 2021 with the descriptors “post-acute Covid-19 syndrome” [Supplementary Concept] OR “Long Covid” [TW].

### SPECT CT reveals cerebral perfusion disorders

The symptoms presented by all patients evoke the same clinical picture made by many authors.^7, 25^ The patients were not aware that their condition was related to Covid. The triad of fatigue, exertional exhaustion and memory impairment seems to be recurrent. Cerebral damage seems to dominate either by cortical damage (brain fog, memory loss, anomia, hallucination, abnormal movement, unexpected paresthesias), or by bulbar damage (anosmia, dysgeusia, orthostatic disorder), although in anosmia, both central and peripheral alterations could be demonstrated.^64^ No patients have contributory NMR images though it can reveal cerebral microvascular lesions in severe Covid-19.^65, 66^

Reviewing the literature on visible alterations on PET scans,^21, 20, 67, 22^ the clinical severity and functional impairment led to replace this technique with brain SPECT CT. PET Scans are unavailable in the first line of care in Belgium and not reimbursed by the Belgian national insurance provider. SPECT CT were therefore ordered in fourteen patients judged severely affected (3 or 4 on the DUSOI /WONCA) and reporting significant functional impairment (score more than 20 in total of the 6 COOP / WONCA charts). Cerebral perfusion changes are visible in all fourteen patients. The images of five of those patients are reproduced here and the lesions found are consistent with the severity of the problem experienced by these patients.

Brain SPECT as a diagnostic tool for demonstration of brain damage was useful in each suspected case. Two severely affected patients each had a follow -up SPECT CT after three months. The SPECT showed a clear improvement in each case, parallel to the clinical improvement of the condition. It should be noted that the improvement in metabolism in the Long Covid at six months has already been shown by the PET Scan.23 This method could thus perhaps also be useful for the follow -up of the most severe cases.

### Default of perfusion and hypercoagulation looks central to Long Covid pathophysiology

The cases presented here are not numerous and were all discovered by the same experimenters in the care setting. But every time the severity of the case and the cognitive disorders are in the foreground, the SPECT shows perfusion abnormalities (14 abnormalities on 14 ordered). These fourteen patients have therefore an encephalopathy by default of perfusion. Not all patients have been ordered a SPECT but the clinical pictures are so similar that one can hardy evoke an another hypothesis to explain the symptoms of the other patients.

Several studies are highlighting the perfusion default as one of the pathophysiological explanation of the clinical symptoms. Mejia et al. suggests a deleterious effect of SARS-CoV-2 infection on systemic vascular endothelial function.^68^ Hohberger et all showed an impaired capillary microcirculation in the macula and peripapillary region.^69^ Fogarty et al. showed that persistent endothelial cell activation may be important in modulating the ongoing pro-coagulant effects in convalescent Covid-19 patients and thus contribute to the pathogenesis underlying the Covid-19 syndrome.^70^ Hypercoagulation, vascular complications^71^ and microclots formation in Covid-19 have already been identified^72^. Long Covid is accompanied by increased levels of antiplasmin,^73^ and pericytes, the multipotent parietal cells of capillaries, may play an important role in microvascular Long Covid alterations^74^

These lesions can be compared to the skin lesions that accompany Covid and Long Covid, especially in young subjects, with a duration of 7 to 150 days.^75^ According to Mehta et al. these lesions clinically resemble vasculopathy, with microvascular abnormalities observed with nail capillaroscopy.^76^ This is an argument for making the analogy between Long Covid and an autoimmune damage or disorder that affects vascular endothelial surfaces. The question arises as to whether the cutaneous vascular lesions in the Covid are similar to the cerebral endothelial lesions inducing the perfusion disorder in the Long Covid. Furthermore, as the skin lesions heal without consequences, one can assume that the cerebrovascular lesions found will evolve in the same way.

Chest pain on exertion could also be attributable to microvascular lesions. Singh and all show that the impaired systemic oxygen extraction observed in exhausted Long Covid patients is attributed primarily to reduced oxygen diffusion in the peripheral microcirculation.^77^ According to an observational study by Camazon et al, coronary microvascular ischemia is the underlying mechanism of persistent chest pain.^78^ The perfusion abnormalities, cognitive and memory impairment could be explained by the expression of ACE2 in the brain stem and other receptors in the cortex by the vascular wall, which makes them vulnerable to the virus.^79,80^ The brainstem regulates respiratory, cardiovascular, gastrointestinal and neurological processes and has a relatively high expression of ACE2 receptors compared to other brain regions.^81^ A perfusional disorder could then explain hyperventilation, abdominal pain or central anosmia for example.

Central perfusion abnormalities, as seen in the present study, and subsequent hypometabolism could explain the cognitive and memory disorders. One can then understand why a patient of the present study, who does not present thalamic and sub-thalamic alterations while the cortical perfusion is very disturbed, presents an intense fatigue, brain fog and effort exhaustion but, surprisingly, no memory disorder.

However, the mechanism of action of the virus on the brain is subject to many hypotheses. The altered mental status may be due to encephalopathy caused by a systemic disease or encephalitis directly caused by the SARS-CoV-2 virus itself.^79^

The reproducibility of the SPECT technique in Long Covid deserve to be studied, as well as its comparison with PET scan. Moreover, SPECT is not an expensive examination, invoiced at 222€ (,250$) to the Belgian national insurance provider.

The environmental impact of labeled Technetium (Tc-99m) must also be taken into account. Although Tc-99m used in medical diagnostics has a short half-life of six hours and does not remain in the body,^82^ its main disadvantages are its high cost and the production of large quantities of highly radioactive waste during its manufacture.^83^ The practitioner requesting such a review should be aware of the environmental impact of the health-related activity.^84^

### Impact of imaging diagnosis on patients’ experiences

Knowing with certainty that the patients’ experiences are not invented but correspond to clear and visible lesions was in each case a shock for the fourteen patients whose SPECT showed altered cerebral perfusion. Despite the anxiety that comes with the diagnosis, the patients are reassured to know that they “*are not crazy* “, that they “*knew that there was something*”, and that “*their family, their employer will finally believe them*”. In other patients, in whom the diagnosis cannot be established with as much precision, it is the clinician’s gradual knowledge and understanding of the clinical picture that will reassure the patient.

This young girl, a passionate gymnast who was called a lazy teenager by her teacher, can finally explain why she could no longer make any effort and why it took her more than 6 months to be at 80% of her capacity again. This worker, who had to stop driving to sleep during his working hours, was reassured by the letter of explanation sent to the occupational physician.The patient mentioned in case 3, now knows from the SPECT images that the previously unknown condition she experienced for more than six months is not a mental disorder. Two patients are comforted by a follow-up SPECT, performed three months after the first one, which shows a clear improvement that is consistent with the clinical picture.

### Persistent diagnostic uncertainty in many patients

However, the diagnosis is not yet clear in all patients. There is no PCR in fourteen of them and three patients are seronegative for Covid, which is also described in the literature.^85^ They are all vaccinated except two. It is currently impossible to distinguish between vaccine immunity and natural immunity within patients. There is therefore no proof that these cases can all, without exception, be labeled as Long Covid. This has consequences for three patients, who cannot attribute their condition with certainty to a disease contracted at work. As a consequence, this deprives them of the benefits of the intervention of the Occupational Diseases Fund.

In an attempt to disentangle the lack of positive lab tests which has repercussions on the diagnosis, a collaboration was obtained with the immunology department of the Catholic University of Leuven (Institut Rega https://rega.kuleuven.be/) and the Karolinska institute (Petter Brodin https://ki.se). In the framework of the European Consortium for Genetic and Immunological Studies on Covid (COVID-HGE consortium https://www.covidhge.com), the patients in this study will benefit from extensive blood tests. The genetic and immunological study tries to understand the occurrence of Long Covid in only some of the Covid patients.^86^ The failure of cytotoxic T and NK cell responses could explain the persistence of the virus (i.e. the virus is alive). It is also possible that Long Covid is the consequence of a dysregulation of the immune system (it would therefore hypothetically be an autoimmune disease).

### Global indicators of severity and functional status

Our indicators of severity (DUSOI) and functional status (COOP/WONCA) are specific to general practice. They sometimes show very severe alterations. The DUSOI is an indicator of the severity estimated by the doctor, while the COOP/WONCA charts represent the patient’s opinion of his or her condition. Many publications deal with the impact of Long Covid on the health status of patients, sometimes with similar indicators,^87^ more detailed ones,^88^ or for example some specific to fatigue.^89^

However, our indicators aim to estimate the overall condition of a patient and not the precise impact on a single function. Patients have other intercurrent health problems and the indicators may be influenced by these new elements. For example, a young patient with emotional loss rated her functional status as very impaired and scored high on the COOP/WONCA charts, while the physician’s assessment of the severity of Long Covid was Grade 2, or mild. The physician will therefore need to relativise the use of these indicators.

### Intercurrent diagnoses

In view of the polymorphous clinical picture of this syndrome, it is not surprising that emergency room colleagues or specialists have put forward diagnoses that are as diverse as they are multiple; angor, pulmonary embolism, hyperventilation, fibromyalgia, traumatic shock, anxiety attacks and post-traumatic stress syndrome, have all been evoked either to exclude them or as peremptory diagnoses that are destabilizing for the patient.

Whether in the emergency room or in with specialists, the patient’s vision is instantaneous and limited to the present moment. The family doctor has a long-term, repetitive, horizontal view. The formulation of unexpected complaints in previously healthy patients, the appearance of inexplicable symptoms, progressive neurological alterations, all sometimes long after an acute episode of Covid, which was not necessarily hospitalized, provoke a questioning and an uneasiness. The strength of longitudinality and relational continuity, defined as “*a continuous therapeutic relationship between a patient and one or more providers*”,^90^ allows the patient to express a growing dismay in the face of an illness that is only getting worse. The long-term doctor-patient relationship allows the co-construction of a reassuring, albeit cruel, diagnostic reality.

### An empathetic therapeutic approach

The announcement of the diagnosis is in itself a therapeutic act. It also makes it possible to set up what seems to be the most coherent long-term therapy, i.e. neurological and physical revalidation. The concept of neuroplasticity, known in other cerebral pathologies,^91^ can be used here to encourage the patient to revalidate his memory. In the Netherlands, a two-arm multi-centric randomized controlled trial (RCT) showed that a comprehensive revalidation program called “Fit after covid” significantly reduces fatigue.^8^

Whether through exercises such as those found in journals like “Brain Sport”, or through the patient’s use of cognitive exercise applications for smartphones, patients must be encouraged to slowly and gradually regain lost ground. One could of course wish for neurocognitive revalidation by specialists in the field, but there are not enough revalidation centers in Belgium.

Getting the body back into action, despite breathing difficulties and pain, is absolutely essential while respecting the limits of each patient. Physiotherapy also has an important role to play, but the Belgian social security system only allows for 18 sessions per condition. Moreover, patients, badly affected by an unknown long-term condition that is damaging their body and mind, need psychological care, which in Belgium has recently been made possible by the establishment of networks of front-line psychologists who are financially accessible. The well-being approach, through massage sessions or thalassotherapy, is not reimbursed by the Belgian social security.

Finally, and this brings us to the crux of the matter, patients who had a job have either lost it or are dependent on social security and only receive 60% of their salary in Belgium. Some are below the poverty line and their distress is great. Social security must be provided to them by adjusting replacement wages. It is necessary that the Belgian state takes into account these patients with this new condition and develops a specific policy of long term support as in the Netherlands^92^ since the year 2020 or in the United Kingdom, which has developed a two-year Long Covid action plan^93^

### What about drug therapy ?

To date, no drug has been shown to be of proven value in the treatment of Long Covid, although many substances are proposed as symptomatic treatments.^94^ Considering the known coagulation disorder in Covid^95^ and the low vascular perfusion seen in Long Covid, one could imagine giving aspirin at a low dose to limit the capacity to make micro-thrombi at the level of the damaged vessels as is done in Covid in high doses.^96^ In a non peer reviewed publication Pretorius and all claim a subset of 24 patients was treated with one month of dual antiplatelet therapy (DAPT) (Clopidogrel 75mg/Aspirin 75mg) once a day, as well as a direct oral anticoagulant (DOAC) (Apixiban) mg twice a day. Each patient reported that their main symptoms were resolved and fatigue as the main symptom was relieved.^97^

Robbins et al. describe a statistically and clinically significant effect often sessions of hyperbaric oxygen therapy on the global fatigue score.^98^ The analogy with the treatment of cluster headaches with normobaric oxygen^99^ could make this therapy, cheap and safe, considered in Long Covid.

In addition, a meta-analysis on the use of plasmapheresis in acute Covid, in mostly intubated patients, showed no effect on mortality but had a positive effect on oxygen partial pressure.^100^ Some practitioners propose it in Long Covid but no publication on this subject has been found.

And last but certainly not least, Hohberger et all. open a therapeutic door by successfully improving a patient through the use of a anti-autoantibody synthetic monoclonal antibody (Aptamer).^69^

## 4.0.1 Conclusion

This observational study is of course limited. Examinations are carried out if they are beneficial to the patient. A practitioner would not ask for SPECT without justification. The unforeseeable is unfortunately not predictable and the information that will prove decisive cannot always be identified beforehand. In his seminal paper on problem solving in general practice, Yan McWhinney wrote that “*the disease presented to family doctors is often in an unorganised state* “.^101^ The practitioner then confronts the information provided by the patient with his or her knowledge and constructs a frame of reference acceptable to him and to the patient.

In this case, there is no pre-organised framework since the condition is new, undescribed and not part of the stock of knowledge accumulated to date. It is therefore the doctor’s ability to listen to the patient, to be surprised and to think about a new situation that will be central to the process of uncovering the new condition. The immense availability of scientific knowledge since the advent of the internet can be used to understand what is going on, to seek the way forward and to assure the patient that his or her case, even if misunderstood, is taken very seriously.

Nevertheless, this study raises more questions than answers. Clinical decisions and methods of patient care should be based on controlled experiments and not on intuition. However, the knowledge of practitioners can also contribute to this and be studied, shared and challenged.^102^ As so well stated by Nisreen A Alwan, *“research is desperately needed to understand Long Covid”* while *“The road to properly addressing Long Covid is long and must be traveled with humility, open mindedness, compassion, and scientific rigor*.*”*^103^

In conclusion, we hope that this contribution will draw the attention of our colleagues to the difficulties encountered by our patients and to the importance of careful listening and monitoring of Long Covid patients. It should be borne in mind that no observation should be considered too small to be valid, and that observations are of little value if they are not shared with peers and subjected to careful analysis. The planned participation in a European multi-center study in genetics and immunology will probably provide some answers and further questions.

## Data Availability

The study was conducted with data from the daily practice of family medicine. The data, carefully de-identified skipping also age, gender, co-morbidities and time course, are available on https://tinyurl.com/tablelongcovid

## Acronym List

APHP: Assistance Publique - Hôpitaux de Paris
COOP Charts: Charts for Primary care Practice ; Cooperative
CT Scan: Computed Tomography
DUSOI: Duke Severity of Illness Score
ECD ED-99m: 99mTc-ethyl cysteinate dimer
EBM: Evidence Based Medicine
EMR: Electronic Medical Record
FP: Family Physician
GP: General practionner
ID: Identity
ICPC-2: International Classification of Primary Care, second version
ISPPC: Intercommunale de Santé Publique du pays de Charleroi
LC: Long Covid
NWR: Nuclear Magnetic Resonance
MeSH: Medical Subject Heading
*PET* Scan: Positron Emission Tomography Computed Tomography
PCR: Polymerase Chain Reaction
SPECT: Single Photon Emission Computed Tomography
WHO: World Health Organization
WONCA: World Organization of Family Doctors

## Ethics

Patients, whose medical records are managed by doctors in a contractual framework, have expressly given their written consent to the use and publication of their personal data in an anonymous manner. The ethics committee of the University Hospital of Liege, Belgium, gave its full approval to this study under the number 2022/23.

## About the authors

Dr Jamoulle, MD, PhD has been a family physician since 1974. He is also a associate researcher at the information sciences department of HEC Liège Management School (University of Liège). Ms. Gisèle Kazeneza-Mugisha is a third year medical student at the University of Mons and was an observer at the consultation of general practice in 2021. Dr. Zayane, MD, has been a family physician since 2017. He works in partnership with Dr. Jamoulle at the Centre Médical Janson, a general practice setting located in Charleroi, Belgium

## Publishing techniques

This text was composed in a shared LATEX format on Overleaf (https://www.overleaf.com/). The Google sheets function was used to edit the summary table of patient data. The bibliography is maintained on Endnote (https://endnote.com/).

The initial text was written in French, translated into English and Spanish by Deepl (https://www.deepl.com/), edited in English by Nora Jamoulle (Brussels, Belgium) and in Spanish by Ricardo La Valle (Buenos Ayres, Argentina) whom we thank wholeheartedly.

## Acknowledgements

Our thanks go to Dr. Salima Bouazza and Dr. Nathalie Mahy, specialist in nuclear medicine at the Vésale Hospital, ISPPC, Belgium, for their availability and their very clear explanations, as well as to Dr. Tatiana Besse, senior research clinician, Brugmann Hospital, Belgium, for her constant scientific support.

## Citation

Jamoulle M, Kazeneza-Mugisha G, Zayane A.Descriptive and narrative study of Long Covid cases in general practice and diagnostic value of single photon emission computed tomography. Clinical research report. Department of General Practice. University of Liège, Belgium. Updated February 2022 ; 36p.

## Footnotes

a. Family doctor and general practitioner are equivalent. The term “general” refers to the extension of the field, the term “family” indicates that he is a doctor of the people.

